# Effectiveness of BNT162b2 XBB Vaccine against XBB and JN.1 Sub-lineages

**DOI:** 10.1101/2024.05.04.24306875

**Authors:** Sara Y. Tartof, Jeff M. Slezak, Laura Puzniak, Timothy B. Frankland, Bradley K. Ackerson, Luis Jodar, John M. McLaughlin

**Affiliations:** Kaiser Permanente Southern California Department of Research & Evaluation, Pasadena, CA, USA; Pfizer Inc., Collegeville, PA, USA; Kaiser Permanente Hawaii Center for Integrated Health Care Research, Honolulu, HI; Southern California Permanente Medical Group, Harbor City, CA, USA

## Abstract

In this Brief Report we provide updated results (October 11, 2023 through February 29, 2024) from our previously conducted test-negative case-control study in Kaiser Permanente Southern California to evaluate sub-lineage-specific effectiveness of the BNT162b2 XBB1.5-adapted vaccine. Results suggest that XBB1.5-adapted vaccines may have reduced effectiveness against JN.1 versus XBB sub-lineages.

## Introduction

JN.1 was first identified in August 2023 and became the predominant SARS-CoV-2 sub-lineage globally by the end of December 2023.^1^ Despite early signs of potential immune evasion,^2,3^ only a few studies in isolated populations^4-6^ have assessed real-world effectiveness of XBB1.5-adapted COVID-19 vaccines against JN.1-related endpoints. We assessed effectiveness of BNT162b2 XBB1.5-adapted vaccine (Pfizer-BioNTech 2023–2024 formulation; hereafter referred to as BNT162b2 XBB vaccine) against XBB and JN.1 sub-lineages in a large, diverse US healthcare system.

## Methods

We updated our previous test-negative case-control BNT162b2 XBB vaccine effectiveness (VE) analysis^7^ to (i) include a longer study period (October 10, 2023 through February 29, 2024 [instead of December 10, 2023]) to allow for additional JN.1 cases, and (ii) stratify VE estimates by time since vaccination and SARS-CoV-2 sub-lineage (i.e., likely XBB- or JN.1-related). Otherwise, selection criteria were identical to our prior analysis.^7^ Briefly, patients were ≥18 years of age who were diagnosed with acute respiratory infection (ARI; **Table S1**) and tested for SARS-CoV-2 using polymerase chain reaction (PCR) during a hospital admission or emergency department (ED) or urgent care (UC) encounter at Kaiser Permanente Southern California during the study period. Cases were those with a positive SARS-CoV-2 PCR test and controls tested negative and had no evidence of a positive SARS-CoV-2 test in the prior 90 days. Likely SARS-CoV-2 strain was determined based on a hierarchy of available information including: (i) whole genome sequencing (WGS), (ii) spike gene target failure (SGTF), or (iii) periods of >80% sub-lineage predominance based on WGS data from US Health and Human Services Region 9.^1^ Odds of receipt of a BNT162b2 XBB vaccine were compared to the odds of not receiving XBB vaccine of any kind (including previously vaccinated and unvaccinated persons) across cases and controls. Adjusted odds ratios (ORs) and 95% confidence intervals (CIs) were calculated using logistic regression and SAS version 9.4. VE was calculated as 1−OR multiplied by 100% (**Supplementary Appendix**).

## Results

Of 59,058 ARI encounters meeting eligibility criteria, 52,036 met selection criteria (8732 [16.8%] hospital admissions, 43,304 [83.2%] ED/UC encounters; **Figure S1**). Median age was 54 years (interquartile range: 38–72; **Tables S2 and S3** describe participant characteristics by case-control and exposure status, respectively). Overall, 7572/52,036 (14.6%) tested SARS-CoV-2 positive, of which 2475/7572 (32.7%) and 2209/7572 (29.2%) were confirmed as likely XBB and JN.1 sub-lineages, respectively. In total, 6923/52,036 (13.3%) received BNT162b2 XBB vaccine with median time since vaccination of 58 days (range: 15–156). Of 7572 cases and 44,464 controls, 753 (9.9%) and 6170 (13.9%), respectively, received BNT162b2 XBB vaccine. Overall (including all sub-lineages) adjusted BNT162b2 XBB VE was 57% (95% CI: 45–66%) against COVID-19-related hospital admission and 40% (34–45%) against ED/UC visits. Against XBB sub-lineages, VE was 65% (41–79%) for hospitalization and 55% (45–64%) for ED/UC; compared to 54% (33–69%) and 41% (32–49%) against JN.1 sub-lineages, respectively (**Figure 1**).

**Figure 1.**
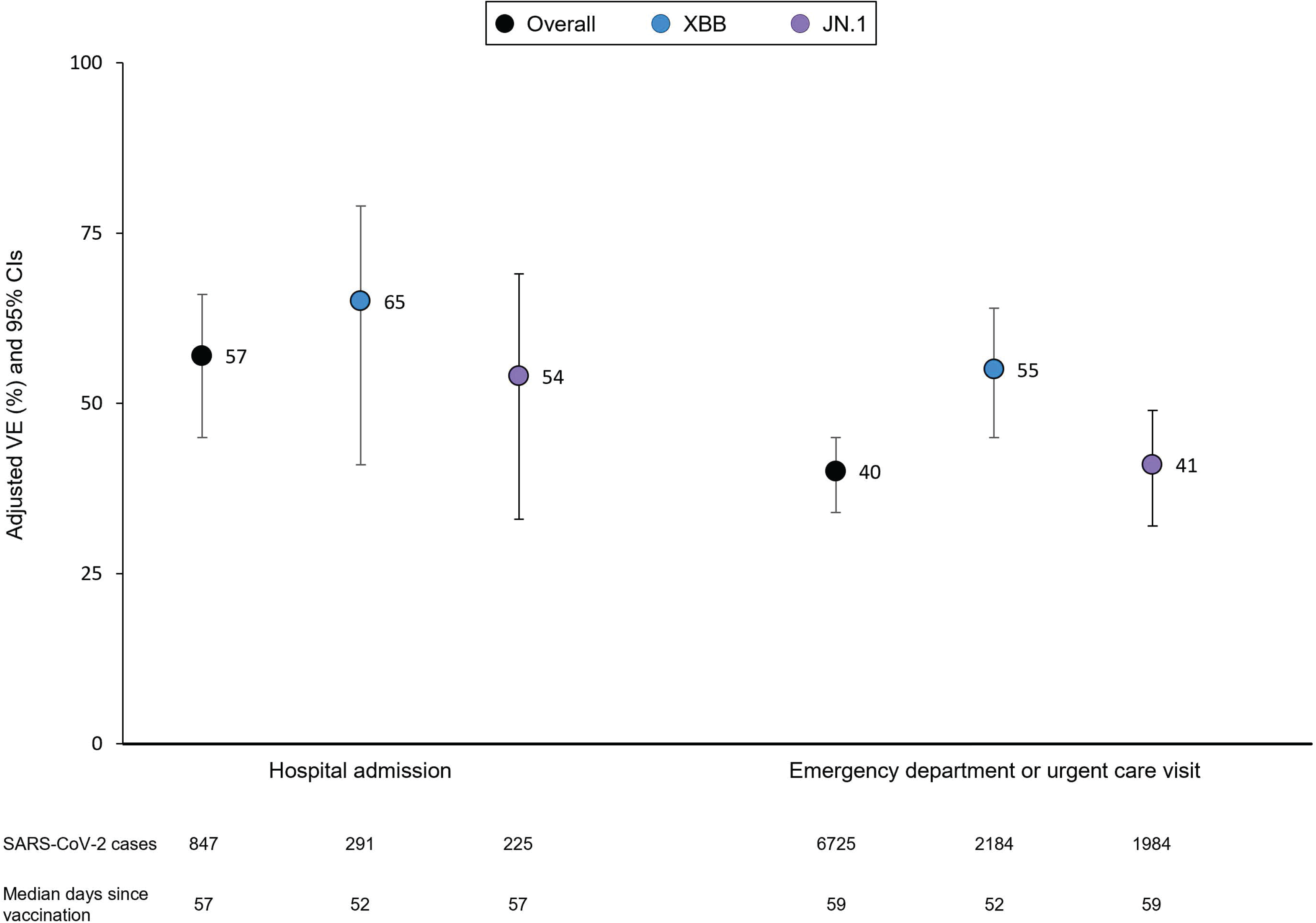
Effectiveness of BNT162b2 XBB vaccine by COVID-19 encounter type and SARS-CoV-2 sub-lineage—October 10, 2023 through February 29, 2024. CI = confidence interval. ED = emergency department; UC = urgent care. Models adjusted for week of encounter, age, sex, self-reported race/ethnicity, BMI, Charlson comorbidity index, prior SARS-CoV-2 infection, and utilization history (flu and pneumococcal vaccination, inpatient, ED, and outpatient encounters in prior year.

When stratified by time since vaccination, VE <60 days post-vaccination against XBB sub-lineages was 74% (49–87%) for hospitalization and 59% (48–68%) for ED/UC; whereas VE against JN.1 sub-lineages for the same two outcomes was 50% (15–71%) and 52% (39–61%), respectively. VE ≥60 days post-vaccination could not be calculated for XBB-related hospitalization due to small sample size, but was 39% (10–59%) against XBB-related ED/UC visits 60–128 days after vaccination. VE was 57% (30–73%) and 34% (22–44%) against JN.1-related hospitalization and ED/UC visits 60–156 days after vaccination, respectively (**Figure 2**).

**Figure 2.**
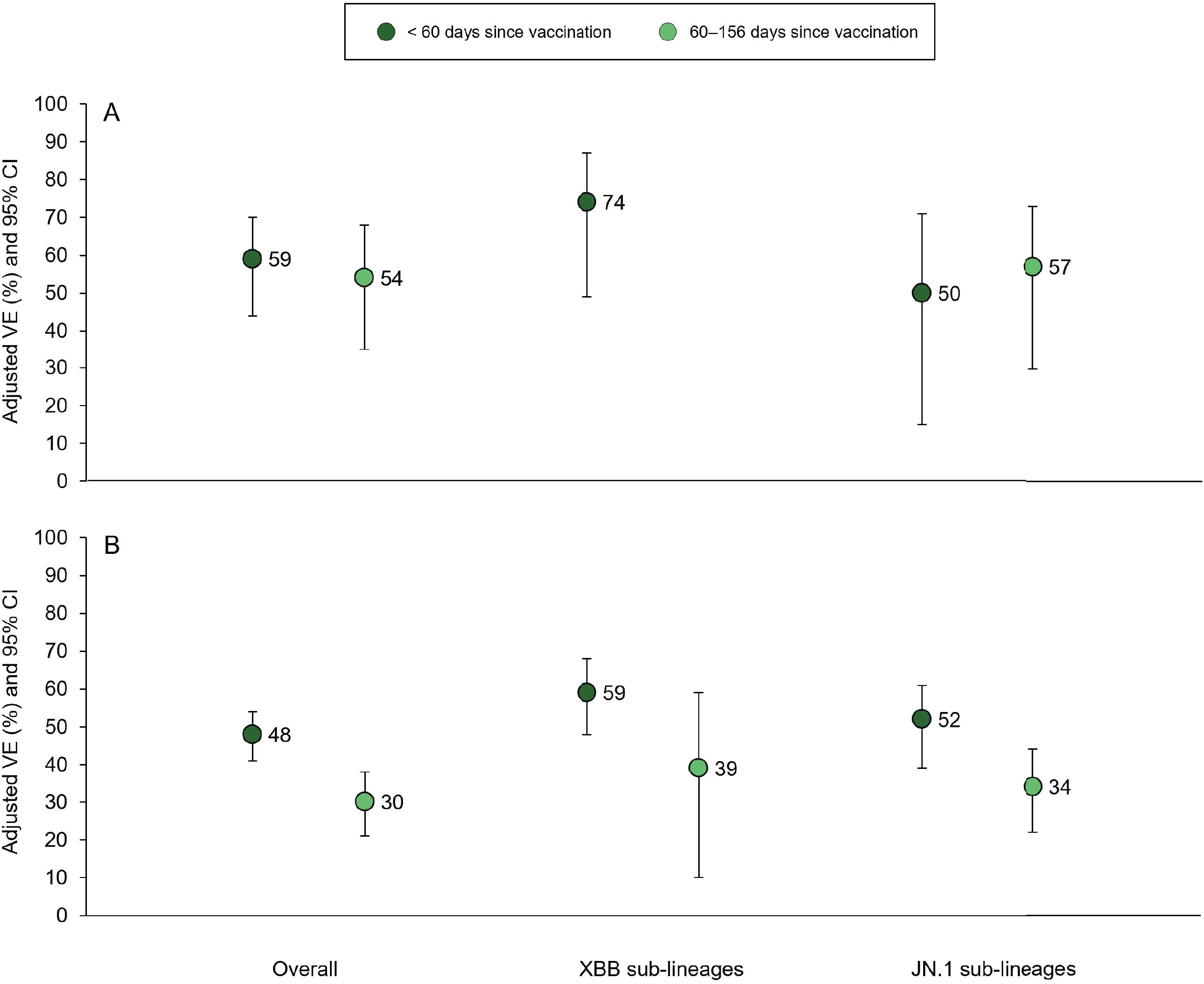
Effectiveness of BNT162b2 XBB vaccine by COVID-19 encounter type, time since vaccination, and SARS-CoV-2 sub-lineage—October 10, 2023 through February 29, 2024. (A) Effectiveness against hospital admission. (B) Effectiveness against ED/UC visits. VE ≥2 months after vaccination could not be calculated for XBB sub-lineages due to small sample size. CI = confidence interval. ED = emergency department; UC = urgent care. Models adjusted for week of encounter, age, sex, self-reported race/ethnicity, BMI, Charlson comorbidity index, prior SARS-CoV-2 infection, and utilization history (flu and pneumococcal vaccination, inpatient, ED, and outpatient encounters in prior year.

## Discussion

In this test-negative case-control study, BNT162b2 XBB VE point estimates were 55–65% against XBB-related outcomes but appeared lower against JN.1 sub-lineages (41–54%), albeit with overlapping CIs. These findings persisted after accounting for time since vaccination.

Consistent with prior reports,^8,9^ VE was highest (>75%) against XBB-related hospitalization within two months of vaccine receipt, a period when XBB1.5-adapted vaccines were most closely matched to circulating XBB strains.^1^ Our observational design has limitations that have been previously described,^7,10,11^ and it is possible that we misclassified SARS-CoV-2 sub-lineages in instances where we relied on SGTF or variant periods. Despite these potential limitations, our results, combined with other early reports,^4-6^ suggest that XBB vaccines likely have reduced effectiveness against JN.1 sub-lineages, which have become predominant globally. Thus, a vaccine strain change for the upcoming 2024–2025 season appears warranted.

## Supporting information

Supplementary Appendix for Effectiveness of BNT162b2 XBB Vaccine against XBB and JN.1 Sub-lineages

## Data Availability

Anonymized data that support the findings of this study may be made available from the investigative team in the following conditions: (1) agreement to collaborate with the study team on all publications, (2) provision of external funding for administrative and investigator time necessary for this collaboration, (3) demonstration that the external investigative team is qualified and has documented evidence of training for human subjects protections, and (4) agreement to abide by the terms outlined in data use agreements between institutions.

## Acknowledgements

TBF and JMS had full access to all the data in the study and takes responsibility for the integrity of the data and the accuracy of the data analysis.

The authors would like to acknowledge Vennis Hong, MPH; Fagen Xie, PhD; Harpreet Takhar, MPH; and Sarah Simmons, MPH from KPSC Department of Research & Evaluation, who received support for their effort paid directly to KPSC. We also thank Joann M. Zamparo, MPH who is an employee of Pfizer Inc and did not receive compensation beyond her salary, for her support of this project.

LJ, LP, and JMM are employees of and hold stock and/or stock options in Pfizer Inc. SYT, TF, JMS, and BA received research support from Pfizer during the conduct of this study that was paid directly to KPSC. BA received research support for work unrelated to this study provided by Pfizer, Moderna, Dynavax, Seqirus, GlaxoSmithKline and Genentech. JMS received research support from ALK, Inc., Dynavax, and Novavax for work unrelated to this study. TBF previously owned stock in Pfizer Inc. SYT received research support from Genentech for work unrelated to this study.

