## Supplementary Appendix for Effectiveness of BNT162b2 XBB Vaccine against XBB and JN.1 Sub-lineages for "Effectiveness of BNT162b2 XBB Vaccine against XBB and JN.1 Sub-lineages"

### Supplementary Methods

*Selection Criteria*

Participants were required to have ≥1 year of health plan membership (allowing for a 31-day gap in membership to account for delays in membership renewal) to determine comorbidities and medical history. Encounters in which the patient had the following were excluded: (1) another positive SARS-CoV-2 test ≤90 days ago, (2) received any type of XBB vaccine other than BNT162b2 XBB vaccine, (3) received a BNT162b2 XBB vaccine ≤2 months after a prior COVID-19 dose, (4) received a BNT162b2 XBB vaccine ≤14 days prior to the encounter, (5) received any other non-XBB booster doses (e.g., BA.4/5 bivalent or wild-type boosters) outside of CDC recommended dosing intervals (recommendations were defined as receipt of any mRNA BA4/.5 bivalent dose between August 31, 2022 and September 11, 2023 with ≥8 weeks [≥56 days] since their most recent dose of original wild-type COVID-19 mRNA vaccine received with a minimal required interval of ≥28 days between a second and subsequent wild-type dose), (6) received nirmatrelvir/ritonavir or any other COVID-19 outpatient antiviral or monoclonal antibody (i.e., molnupiravir, remdesivir, bebtelovimab, bamlanivimab, casirivimab, cilgavimab, sotrovimab, tixagevimab) in the 30 days prior to a COVID-19 encounter, or (7) a hospital admission that, despite having an ARI diagnosis with a positive SARS-CoV-2 test, was determined to be likely unrelated to COVID-19 or clearly related to another cause based on medical chart review that was conducted by trained research staff who were blinded to vaccination status and later validated by a blinded physician investigator (BKA). For patients that had multiple encounters, we included only the first encounter to maintain independence of outcome events.

*Outcomes*

Hospital admission and ED/UC outcomes were mutually exclusive. SARS-CoV-2 PCR tests among cases and controls were restricted to those administered ≤14 days prior to the initial ARI encounter through ≤3 days after the encounter.

*Exposures*

All members were eligible for COVID-19 vaccines at no cost based on FDA authorized or approved indications. KPSC EHRs captured all vaccinations administered within the health system. Records were supplemented with vaccine administration data from the California Immunization Registry, to which all healthcare providers are required by law to report COVID-19 vaccinations within 24 hours. To be considered vaccinated, the dose had to occur >14 days before testing for SARS-CoV-2.

*Identification of SARS-CoV-2 Sub-lineage*

Likely variant sub-lineage was determined in a hierarchical fashion. If available, we first used whole-genome sequencing (WGS) results to identify variant sub-lineage. Viral sequencing was performed by Helix (San Mateo, CA, USA) using a hybridization-capture based assay (Twist Biosciences, San Francisco, CA, USA) and short-read genome sequencing technology (Illumina, San Diego, CA, USA).[1] Lineages were assigned using pangolin version 4.3.1. If WGS data were not available, we used S-gene target failure (SGTF) results as measured by the TaqPath COVID-19 Combo Kit (ThermoFisher, Waltham, MA, USA) to distinguish between XBB sub-lineages (SGTF negative) and JN.1 sub-lineages (SGTF positive). Finally, for positive SARS-CoV-2 tests where variant sub-lineage could not be determined using WGS or SGTF, we assigned sub-lineage using calendar time. Specifically, we identified periods when XBB or JN.1 sub-lineages exceeded roughly 80% of all sequenced strains in US Health and Human Services Region 9 (which includes the State of California).[2] Accordingly, cases were classified as XBB sub-lineages from October 10 through December 9, 2023; and as JN.1 sub-lineages from January 20 through February 29, 2024.[2] Overall, 1604/2475 (64.8%), 215/2475 (8.7%), and 656/2475 (26.5%) of XBB sub-lineages were determined by WGS, SGTF, or calendar time (i.e., variant predominant period), respectively. The same percentages for JN.1 determination were 1189/2209 (53.8%), 211/2209 (9.6%), and 809/2209 (36.6%), respectively. For XBB sub-lineage analyses, test-negative controls were identified between October 10, 2023 through February 2, 2024 (date of the last WGS-confirmed XBB sub-lineage case at KPSC). For JN.1 sub-lineage analyses, test-negative controls were identified between October 30, 2023 (date of the first WGS-confirmed JN.1 sub-lineage case at KPSC) through February 29, 2024.

*Statistical Analysis*

Odds ratios (ORs) and 95% confidence intervals (CIs) calculated using the Wald method were derived from multivariable logistic regression models that included week of encounter, age (18−49, 50−64, and ≥65 years), sex (female, male), self-reported race/ethnicity (non-Hispanic Asian/Pacific Islander, non-Hispanic African-American/Black, Hispanic/Latinx, non-Hispanic White, other [including individuals who identified as American Indian or multiple or other race and ethnicity], and unknown), body mass index (<18.5, 18.5−24.9, 25.0−29.9, 30.0−34.9, ≥35.0 $kg/m^2$, and unknown), Charlson risk score (0, 1, 2, 3, and ≥4), receipt of influenza vaccine in the year before admission (yes or no), receipt of pneumococcal vaccine in the 5 years before admission (yes or no), health-care utilization in the year before admission (i.e., number of hospital admissions and ED or outpatient visits), and documentation of previous SARS-CoV-2 infection confirmed by PCR or antigen test (ever *vs* never) for pre-delta, delta, and omicron periods. Missing values were treated as separate categories for all variables in all analyses.

Analyses were done separately for ARI-associated hospital admissions and ED/UC encounters, and separately by likely variant sub-lineage as defined above. Vaccine effectiveness (VE) was calculated as 1−OR multiplied by 100%. This study was approved by the KPSC institutional review board which waived the requirement for informed consent.

**Table S1.** Acute Respiratory Infection Codes, October 10, 2023 through December 10, 2023

| | |
|---|---|
| A48.1 | LEGIONNAIRES' DISEASE |
| B34.2 | CORONAVIRUS INFECTION, UNSPECIFIED |
| B44.0 | INVASIVE PULMONARY ASPERGILLOSIS |
| B97.29 | OTH CORONAVIRUS AS THE CAUSE OF DISEASES CLASSD ELSWHR |
| J00 | ACUTE NASOPHARYNGITIS (COMMON COLD) |
| J01.00 | ACUTE MAXILLARY SINUSITIS, UNSPECIFIED |
| J01.10 | ACUTE FRONTAL SINUSITIS, UNSPECIFIED |
| J01.20 | ACUTE ETHMOIDAL SINUSITIS, UNSPECIFIED |
| J01.30 | ACUTE SPHENOIDAL SINUSITIS, UNSPECIFIED |
| J01.40 | ACUTE PANSINUSITIS, UNSPECIFIED |
| J01.80 | OTHER ACUTE SINUSITIS |
| J01.90 | ACUTE SINUSITIS, UNSPECIFIED |
| J02.0 | STREPTOCOCCAL PHARYNGITIS |
| J02.8 | ACUTE PHARYNGITIS DUE TO OTHER SPECIFIED ORGANISMS |
| J02.9 | ACUTE PHARYNGITIS, UNSPECIFIED |
| J03.00 | ACUTE STREPTOCOCCAL TONSILLITIS, UNSPECIFIED |
| J03.90 | ACUTE TONSILLITIS, UNSPECIFIED |
| J04.0 | ACUTE LARYNGITIS |
| J04.10 | ACUTE TRACHEITIS WITHOUT OBSTRUCTION |
| J05.0 | ACUTE OBSTRUCTIVE LARYNGITIS (CROUP) |
| J05.10 | ACUTE EPIGLOTTITIS WITHOUT OBSTRUCTION |
| J06.0 | ACUTE LARYNGOPHARYNGITIS |
| J06.9 | ACUTE UPPER RESPIRATORY INFECTION, UNSPECIFIED |
| J09.X1 | INFLUENZA DUE TO IDENT NOVEL INFLUENZA A VIRUS W PNEUMONIA |
| J09.X2 | FLU DUE TO IDENT NOVEL INFLUENZA A VIRUS W OTH RESP MANIFEST |
| J10.00 | FLU DUE TO OTH IDENT FLU VIRUS W UNSP TYPE OF PNEUMONIA |
| J10.01 | FLU DUE TO OTH IDENT FLU VIRUS W SAME OTH IDENT FLU VIRUS PN |
| J10.08 | INFLUENZA DUE TO OTH IDENT INFLUENZA VIRUS W OTH PNEUMONIA |
| J10.1 | FLU DUE TO OTH IDENT INFLUENZA VIRUS W OTH RESP MANIFEST |
| J10.2 | INFLUENZA DUE TO OTH IDENT INFLUENZA VIRUS W GI MANIFEST |
| J11.00 | FLU DUE TO UNIDENTIFIED FLU VIRUS W UNSP TYPE OF PNEUMONIA |
| J11.08 | FLU DUE TO UNIDENTIFIED FLU VIRUS W SPECIFIED PNEUMONIA |
| J11.1 | FLU DUE TO UNIDENTIFIED INFLUENZA VIRUS W OTH RESP MANIFEST |
| J12.1 | RESPIRATORY SYNCYTIAL VIRUS PNEUMONIA |
| J12.2 | PARAINFLUENZA VIRUS PNEUMONIA |
| J12.3 | HUMAN METAPNEUMOVIRUS PNEUMONIA |
| J12.81 | PNEUMONIA DUE TO SARS-ASSOCIATED CORONAVIRUS |
| J12.82 | PNEUMONIA DUE TO CORONAVIRUS DISEASE 2019 |
| J12.89 | OTHER VIRAL PNEUMONIA |
| J12.9 | VIRAL PNEUMONIA, UNSPECIFIED |

| | |
|---|---|
| J13 | PNEUMONIA DUE TO STREPTOCOCCUS PNEUMONIAE |
| J14 | PNEUMONIA DUE TO HEMOPHILUS INFLUENZAE |
| J15.0 | PNEUMONIA DUE TO KLEBSIELLA PNEUMONIAE |
| J15.1 | PNEUMONIA DUE TO PSEUDOMONAS |
| J15.20 | PNEUMONIA DUE TO STAPHYLOCOCCUS, UNSPECIFIED |
| J15.211 | PNEUMONIA DUE TO METHICILLIN SUSCEP STAPH |
| J15.212 | PNEUMONIA DUE TO METHICILLIN RESISTANT STAPHYLOCOCCUS AUREUS |
| J15.4 | PNEUMONIA DUE TO OTHER STREPTOCOCCI |
| J15.5 | PNEUMONIA DUE TO ESCHERICHIA COLI |
| J15.6 | PNEUMONIA DUE TO OTHER AEROBIC GRAM-NEGATIVE BACTERIA |
| J15.7 | PNEUMONIA DUE TO MYCOPLASMA PNEUMONIAE |
| J15.8 | PNEUMONIA DUE TO OTHER SPECIFIED BACTERIA |
| J15.9 | UNSPECIFIED BACTERIAL PNEUMONIA |
| J16.8 | PNEUMONIA DUE TO OTHER SPECIFIED INFECTIOUS ORGANISMS |
| J18.0 | BRONCHOPNEUMONIA, UNSPECIFIED ORGANISM |
| J18.1 | LOBAR PNEUMONIA, UNSPECIFIED ORGANISM |
| J18.8 | OTHER PNEUMONIA, UNSPECIFIED ORGANISM |
| J18.9 | PNEUMONIA, UNSPECIFIED ORGANISM |
| J20.2 | ACUTE BRONCHITIS DUE TO STREPTOCOCCUS |
| J20.5 | ACUTE BRONCHITIS DUE TO RESPIRATORY SYNCYTIAL VIRUS |
| J20.6 | ACUTE BRONCHITIS DUE TO RHINOVIRUS |
| J20.8 | ACUTE BRONCHITIS DUE TO OTHER SPECIFIED ORGANISMS |
| J20.9 | ACUTE BRONCHITIS, UNSPECIFIED |
| J22 | UNSPECIFIED ACUTE LOWER RESPIRATORY INFECTION |
| J39.0 | RETROPHARYNGEAL AND PARAPHARYNGEAL ABSCESS |
| J39.1 | OTHER ABSCESS OF PHARYNX |
| J39.2 | OTHER DISEASES OF PHARYNX |
| J39.8 | OTHER SPECIFIED DISEASES OF UPPER RESPIRATORY TRACT |
| J80 | ACUTE RESPIRATORY DISTRESS SYNDROME |
| J96.00 | ACUTE RESPIRATORY FAILURE, UNSP W HYPOXIA OR HYPERCAPNIA |
| J96.01 | ACUTE RESPIRATORY FAILURE WITH HYPOXIA |
| J96.02 | ACUTE RESPIRATORY FAILURE WITH HYPERCAPNIA |
| J96.10 | CHRONIC RESPIRATORY FAILURE, UNSP W HYPOXIA OR HYPERCAPNIA |
| J96.11 | CHRONIC RESPIRATORY FAILURE WITH HYPOXIA |
| J96.12 | CHRONIC RESPIRATORY FAILURE WITH HYPERCAPNIA |
| J96.20 | ACUTE AND CHR RESP FAILURE, UNSP W HYPOXIA OR HYPERCAPNIA |
| J96.21 | ACUTE AND CHRONIC RESPIRATORY FAILURE WITH HYPOXIA |
| J96.22 | ACUTE AND CHRONIC RESPIRATORY FAILURE WITH HYPERCAPNIA |
| J96.90 | RESPIRATORY FAILURE, UNSP, UNSP W HYPOXIA OR HYPERCAPNIA |
| J96.91 | RESPIRATORY FAILURE, UNSPECIFIED WITH HYPOXIA |
| J96.92 | RESPIRATORY FAILURE, UNSPECIFIED WITH HYPERCAPNIA |
| M35.81 | MULTISYSTEM INFLAMATORY SYNDROME |

| | |
|---|---|
| M35.89 | OTHER SPECIFIED SYSTEMIC INVOLVMENT OF CONNECTIVE TISSUE |
| R05.1 | ACUTE COUGH |
| R05.3 | CHRONIC COUGH |
| R05.4 | COUGH SYNCOPE |
| R05.8 | OTHER SPECIFIED COUGH |
| R05.9 | COUGH, UNSPECIFIED |
| R09.2 | RESPIRATORY ARREST |
| R50.9 | FEVER, UNSPECIFIED |
| U07.1 | COVID-19 |

**Table S2. Study population characteristics by case-control status**

| *Characteristic* | *XBB cases* | *JN.1 cases* | *Other or unknown cases* | *Controls* | *Total* |
|---|---|---|---|---|---|
| ***Total N*** | 2475 | 2209 | 2888 | 44464 | 52036 |
| ***Prior COVID-19 vaccination*** | | | | | |
| *Unvaccinated* | 295 (11.9%) | 205 (9.3%) | 357 (12.4%) | 5408 (12.2%) | 6265 (12%) |
| *1 original wild-type dose and no BA.4/5 bivalent doses* | 70 (2.8%) | 47 (2.1%) | 79 (2.7%) | 1388 (3.1%) | 1584 (3%) |
| *2 original wild-type doses and no BA.4/5 bivalent doses* | 486 (19.6%) | 432 (19.6%) | 605 (20.9%) | 9103 (20.5%) | 10626 (20.4%) |
| *≥3 original wild-type doses and no BA.4/5 bivalent dose* | 923 (37.3%) | 833 (37.7%) | 957 (33.1%) | 14515 (32.6%) | 17228 (33.1%) |
| *≥1 BA.4/5 bivalent doses* | 701 (28.3%) | 692 (31.3%) | 890 (30.8%) | 14050 (31.6%) | 16333 (31.4%) |
| ***Age in years at time of encounter*** | | | | | |
| *18–49* | 884 (35.7%) | 756 (34.2%) | 1150 (39.8%) | 18373 (41.3%) | 21163 (40.7%) |
| *50–64* | 602 (24.3%) | 529 (23.9%) | 631 (21.8%) | 10073 (22.7%) | 11835 (22.7%) |
| *≥65* | 989 (40%) | 924 (41.8%) | 1107 (38.3%) | 16018 (36%) | 19038 (36.6%) |
| ***Sex*** | | | | | |
| *Female* | 1450 (58.6%) | 1258 (56.9%) | 1763 (61%) | 25868 (58.2%) | 30339 (58.3%) |
| *Male* | 1025 (41.4%) | 951 (43.1%) | 1125 (39%) | 18596 (41.8%) | 21697 (41.7%) |
| ***Self-Reported Race / Ethnicity*** | | | | | |
| *Non-Hispanic African American/Black* | 280 (11.3%) | 272 (12.3%) | 369 (12.8%) | 4974 (11.2%) | 5895 (11.3%) |
| *Non-Hispanic Asian/Pacific Islander* | 235 (9.5%) | 242 (11%) | 308 (10.7%) | 4596 (10.3%) | 5381 (10.3%) |
| *Hispanic/Latinx* | 1122 (45.3%) | 1015 (45.9%) | 1285 (44.5%) | 20140 (45.3%) | 23562 (45.3%) |
| *Non-Hispanic White* | 740 (29.9%) | 597 (27%) | 801 (27.7%) | 1805 (4.1%) | 2111 (4.1%) |
| *Other/Unknown** | 98 (4%) | 83 (3.8%) | 125 (4.3%) | 12949 (29.1%) | 15087 (29%) |
| | | | | | **cont.** |

| | | | | | |
|---|---|---|---|---|---|
| ***Encounter type*** | | | | | |
| *Hospital admission* | 291 (11.8%) | 225 (10.2%) | 331 (11.5%) | 7885 (17.7%) | 8732 (16.8%) |
| *Emergency department or urgent care visit* | 2184 (88.2%) | 1984 (89.8%) | 2557 (88.5%) | 36579 (82.3%) | 43304 (83.2%) |
| ***Charlson comorbidity index*** | | | | | |
| *0* | 1073 (43.4%) | 966 (43.7%) | 1310 (45.4%) | 19657 (44.2%) | 23006 (44.2%) |
| *1* | 431 (17.4%) | 400 (18.1%) | 509 (17.6%) | 8022 (18%) | 9362 (18%) |
| *2* | 263 (10.6%) | 220 (10%) | 315 (10.9%) | 4401 (9.9%) | 5199 (10%) |
| *3* | 169 (6.8%) | 164 (7.4%) | 188 (6.5%) | 2825 (6.4%) | 3346 (6.4%) |
| *≥4* | 539 (21.8%) | 459 (20.8%) | 566 (19.6%) | 9559 (21.5%) | 11123 (21.4%) |
| ***Body mass index (BMI; kg/m$^2$)*** | | | | | |
| *Underweight (<18.5)* | 55 (2.2%) | 49 (2.2%) | 66 (2.3%) | 1030 (2.3%) | 1200 (2.3%) |
| *Normal or healthy weight (18.5–24.9)* | 579 (23.4%) | 562 (25.4%) | 696 (24.1%) | 10033 (22.6%) | 11870 (22.8%) |
| *Overweight (25.0–29.9)* | 775 (31.3%) | 688 (31.1%) | 892 (30.9%) | 13139 (29.5%) | 15494 (29.8%) |
| *Obese, class 1 (30.0–34.9)* | 564 (22.8%) | 485 (22%) | 638 (22.1%) | 10134 (22.8%) | 11821 (22.7%) |
| *Obese, class 2-3 (≥35.0)* | 484 (19.6%) | 409 (18.5%) | 573 (19.8%) | 9677 (21.8%) | 11143 (21.4%) |
| *Unknown* | 18 (0.7%) | 16 (0.7%) | 23 (0.8%) | 451 (1%) | 508 (1%) |
| ***Prior documented positive SARS-CoV-2 PCR or antigen tests*** | | | | | |
| *Pre-delta era (March 1, 2020 – June 11, 2021)* | 255 (10.3%) | 190 (8.6%) | 283 (9.8%) | 4641 (10.4%) | 5369 (10.3%) |
| *Delta era (June 12, 2021 – December 20, 2021)* | 56 (2.3%) | 58 (2.6%) | 72 (2.5%) | 1331 (3%) | 1517 (2.9%) |
| *Omicron era (December 21, 2021 – current)* | 515 (20.8%) | 569 (25.8%) | 719 (24.9%) | 12939 (29.1%) | 14742 (28.3%) |
| *No prior documented SARS-CoV-2 test* | 1715 (69.3%) | 1457 (66%) | 1901 (65.8%) | 27236 (61.3%) | 32309 (62.1%) |
| | | | | | **cont.** |

| | | | | | |
|---|---|---|---|---|---|
| ***Healthcare Utilization (counts) in year prior to encounter*** | | | | | |
| *Outpatient Visits* | | | | | |
| *0* | 119 (4.8%) | 93 (4.2%) | 149 (5.2%) | 2637 (5.9%) | 2998 (5.8%) |
| *1* | 148 (6%) | 119 (5.4%) | 141 (4.9%) | 2526 (5.7%) | 2934 (5.6%) |
| *2–4* | 409 (16.5%) | 410 (18.6%) | 533 (18.5%) | 8387 (18.9%) | 9739 (18.7%) |
| *5–9* | 614 (24.8%) | 528 (23.9%) | 730 (25.3%) | 10856 (24.4%) | 12728 (24.5%) |
| *≥10* | 1185 (47.9%) | 1059 (47.9%) | 1335 (46.2%) | 20058 (45.1%) | 23637 (45.4%) |
| *Emergency Department Visits* | | | | | |
| *0* | 1545 (62.4%) | 1412 (63.9%) | 1861 (64.4%) | 27666 (62.2%) | 32484 (62.4%) |
| *1* | 495 (20%) | 427 (19.3%) | 538 (18.6%) | 8624 (19.4%) | 10084 (19.4%) |
| *≥2* | 435 (17.6%) | 370 (16.7%) | 489 (16.9%) | 8174 (18.4%) | 9468 (18.2%) |
| *Hospital Admissions* | | | | | |
| *0* | 2150 (86.9%) | 1938 (87.7%) | 2513 (87%) | 37902 (85.2%) | 44503 (85.5%) |
| *1* | 225 (9.1%) | 204 (9.2%) | 242 (8.4%) | 4073 (9.2%) | 4744 (9.1%) |
| *≥2* | 100 (4%) | 67 (3%) | 133 (4.6%) | 2489 (5.6%) | 2789 (5.4%) |
| ***Received influenza vaccine in year prior to encounter*** | | | | | |
| *No* | 1125 (45.5%) | 901 (40.8%) | 1348 (46.7%) | 20991 (47.2%) | 24365 (46.8%) |
| *Yes* | 1350 (54.5%) | 1308 (59.2%) | 1540 (53.3%) | 23473 (52.8%) | 27671 (53.2%) |
| ***Received pneumococcal vaccine in 5 years prior to encounter*** | | | | | |
| *No* | 1861 (75.2%) | 1660 (75.1%) | 2186 (75.7%) | 33306 (74.9%) | 39013 (75%) |
| *Yes* | 614 (24.8%) | 549 (24.9%) | 702 (24.3%) | 11158 (25.1%) | 13023 (25%) |

**Table S3. Study population characteristics by XBB vaccine receipt.**

| *Characteristic* | *Did not receive an XBB vaccine of any kind* | *Received BNT162b2 XBB vaccine* | *Total* |
|---|---|---|---|
| ***Total N*** | 45113 | 6923 | 52036 |
| ***Prior COVID-19 vaccination*** | | | |
| *Unvaccinated* | 6265 (13.9%) | 0 (0%) | 6265 (12%) |
| *1 original wild-type dose and no BA.4/5 bivalent doses* | 1584 (3.5%) | 0 (0%) | 1584 (3%) |
| *2 original wild-type doses and no BA.4/5 bivalent doses* | 10461 (23.2%) | 165 (2.4%) | 10626 (20.4%) |
| *≥3 original wild-type doses and no BA.4/5 bivalent dose* | 15900 (35.2%) | 1328 (19.2%) | 17228 (33.1%) |
| *≥1 BA.4/5 bivalent doses* | 10903 (24.2%) | 5430 (78.4%) | 16333 (31.4%) |
| ***Age in years at time of encounter*** | | | |
| *18–49* | 19964 (44.3%) | 1199 (17.3%) | 21163 (40.7%) |
| *50–64* | 10449 (23.2%) | 1386 (20%) | 11835 (22.7%) |
| *≥65* | 14700 (32.6%) | 4338 (62.7%) | 19038 (36.6%) |
| ***Sex*** | | | |
| *Female* | 26580 (58.9%) | 3759 (54.3%) | 30339 (58.3%) |
| *Male* | 18533 (41.1%) | 3164 (45.7%) | 21697 (41.7%) |
| ***Self-Reported Race / Ethnicity*** | | | |
| *Non-Hispanic African American/Black* | 5236 (11.6%) | 659 (9.5%) | 5895 (11.3%) |
| *Non-Hispanic Asian/Pacific Islander* | 4380 (9.7%) | 1001 (14.5%) | 5381 (10.3%) |
| *Hispanic/Latinx* | 21295 (47.2%) | 2267 (32.7%) | 23562 (45.3%) |
| *Non-Hispanic White* | 1911 (4.2%) | 200 (2.9%) | 2111 (4.1%) |
| *Other/Unknown** | 12291 (27.2%) | 2796 (40.4%) | 15087 (29%) |
| ***Encounter type*** | | | |
| *Hospital admission* | 7256 (16.1%) | 1476 (21.3%) | 8732 (16.8%) |
| *Emergency department or urgent care visit* | 37857 (83.9%) | 5447 (78.7%) | 43304 (83.2%) |
| | | | **cont.** |

| | | | |
|---|---|---|---|
| ***Charlson comorbidity index*** | | | |
| *0* | 21277 (47.2%) | 1729 (25%) | 23006 (44.2%) |
| *1* | 8140 (18%) | 1222 (17.7%) | 9362 (18%) |
| *2* | 4238 (9.4%) | 961 (13.9%) | 5199 (10%) |
| *3* | 2659 (5.9%) | 687 (9.9%) | 3346 (6.4%) |
| *≥4* | 8799 (19.5%) | 2324 (33.6%) | 11123 (21.4%) |
| ***Body mass index (BMI; kg/m²)*** | | | |
| *Underweight (<18.5)* | 1040 (2.3%) | 160 (2.3%) | 1200 (2.3%) |
| *Normal or healthy weight (18.5–24.9)* | 10117 (22.4%) | 1753 (25.3%) | 11870 (22.8%) |
| *Overweight (25.0–29.9)* | 13277 (29.4%) | 2217 (32%) | 15494 (29.8%) |
| *Obese, class 1 (30.0–34.9)* | 10307 (22.8%) | 1514 (21.9%) | 11821 (22.7%) |
| *Obese, class 2-3 (≥35.0)* | 9880 (21.9%) | 1263 (18.2%) | 11143 (21.4%) |
| *Unknown* | 492 (1.1%) | 16 (0.2%) | 508 (1%) |
| ***Prior documented positive SARS-CoV-2 PCR or antigen tests*** | | | |
| *Pre-delta era (March 1, 2020 – June 11, 2021)* | 4949 (11%) | 420 (6.1%) | 5369 (10.3%) |
| *Delta era (June 12, 2021 – December 20, 2021)* | 1407 (3.1%) | 110 (1.6%) | 1517 (2.9%) |
| *Omicron era (December 21, 2021 – current)* | 12804 (28.4%) | 1938 (28%) | 14742 (28.3%) |
| *No prior documented SARS-CoV-2 test* | 27727 (61.5%) | 4582 (66.2%) | 32309 (62.1%) |
| ***Healthcare Utilization (counts) in year prior to encounter*** | | | |
| *Outpatient Visits* | | | |
| *0* | 2984 (6.6%) | 14 (0.2%) | 2998 (5.8%) |
| *1* | 2815 (6.2%) | 119 (1.7%) | 2934 (5.6%) |
| *2–4* | 9120 (20.2%) | 619 (8.9%) | 9739 (18.7%) |
| *5–9* | 11201 (24.8%) | 1527 (22.1%) | 12728 (24.5%) |
| *≥10* | 18993 (42.1%) | 4644 (67.1%) | 23637 (45.4%) |
| *Emergency Department Visits* | | | |
| *0* | 28337 (62.8%) | 4147 (59.9%) | 32484 (62.4%) |
| *1* | 8754 (19.4%) | 1330 (19.2%) | 10084 (19.4%) |
| *≥2* | 8022 (17.8%) | 1446 (20.9%) | 9468 (18.2%) |
| | | | **cont.** |

| | | | |
|---|---|---|---|
| *Hospital Admissions* | | | |
| *0* | 38762 (85.9%) | 5741 (82.9%) | 44503 (85.5%) |
| *1* | 3986 (8.8%) | 758 (10.9%) | 4744 (9.1%) |
| *≥2* | 2365 (5.2%) | 424 (6.1%) | 2789 (5.4%) |
| ***Received influenza vaccine in year prior to encounter*** | | | |
| *No* | 24093 (53.4%) | 272 (3.9%) | 24365 (46.8%) |
| *Yes* | 21020 (46.6%) | 6651 (96.1%) | 27671 (53.2%) |
| ***Received pneumococcal vaccine in 5 years prior to encounter*** | | | |
| *No* | 34483 (76.4%) | 4530 (65.4%) | 39013 (75%) |
| *Yes* | 10630 (23.6%) | 2393 (34.6%) | 13023 (25%) |

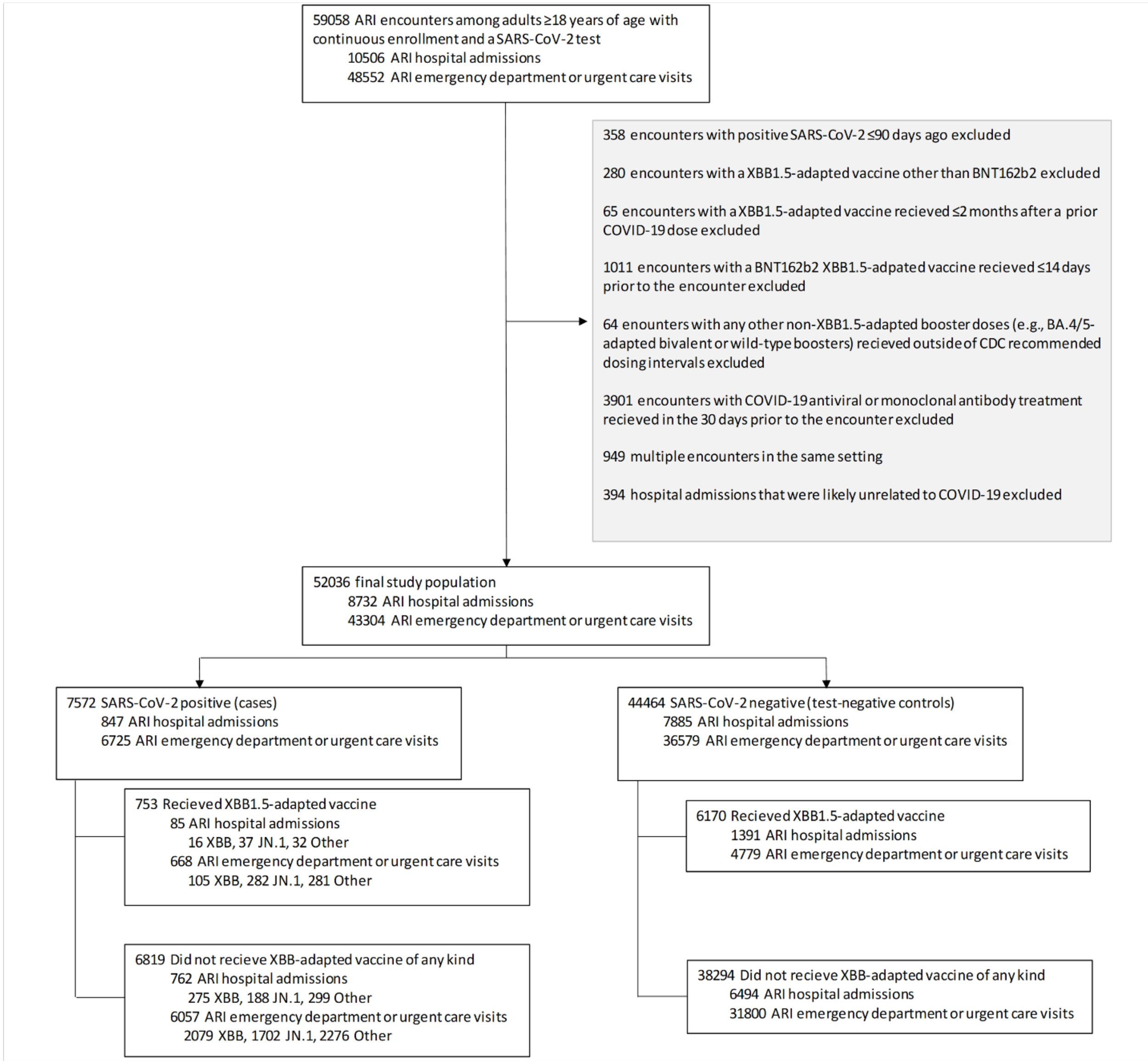

**Figure S1. Selection criteria**

ARI = acute respiratory infection. CDC = Centers for Disease Control and Prevention.
